# Network-based Plasma Proteomics Reveals Molecular Overlap Between Physical Activity and Dementia Risk

**DOI:** 10.1101/2025.03.07.25323587

**Authors:** Rowan Saloner, Emily W. Paolillo, Anna M. VandeBunte, Claire J. Cadwallader, Coty Chen, Brian T. Steffen, David A. Bennett, Bradley F. Boeve, Howard J. Rosen, Adam L. Boxer, Joel H. Kramer, Kaitlin B. Casaletto

## Abstract

Physical activity (PA) is linked to lower dementia risk, but molecular pathways underpinning PA-related dementia risk are poorly understood. We conducted plasma proteomics (SomaScan v4.1) and 30-day Fitbit-based PA monitoring (average daily step count) in 65 cognitively unimpaired older adults from the UCSF BrANCH cohort. Differential regression and network analyses identified PA plasma proteomic signatures tied to extracellular matrix (ECM), immune response, and lipid metabolism. Protein module M12 ECM/neurodevelopment positively correlated with PA in BrANCH and external cohorts, inversely predicted cognitive aging outcomes in BrANCH, and decreased across multiple neurodegenerative conditions. M12 was enriched for proteins from Alzheimer’s disease (AD) risk genes and antemortem plasma abundance of ANTXR2, an M12 ‘hub’ protein, forecasted longitudinal cognitive decline and postmortem brain tissue protein signatures of AD cognitive resilience in the ROSMAP cohort. Our integrated analysis across six proteomic datasets identified blood-detectable molecular signatures of PA and neurodegenerative disease, including ECM-related proteins (e.g., ANTXR2) that may represent key molecular targets for dementia prevention.

## INTRODUCTION

Dementia represents a significant global health challenge, with limited therapeutic options currently available (*1*). Physical activity (PA) is a modifiable lifestyle factor that has consistently demonstrated neuroprotective effects, including associations with lower incidence of dementia (*2*), less cortical atrophy (*3, 4*) and slower neurodegenerative trajectories in individuals with sporadic and even autosomal dominant dementia pathophysiology (*5–7*). Despite these promising findings, the molecular pathways underlying relationships between PA and neurodegeneration remain poorly understood. Identifying biological signatures of PA will enable stratification of individuals who may benefit most from PA interventions and could uncover novel therapeutic targets for dementia prevention.

High-throughput proteomic platforms allow for quantification of thousands of proteins in biofluid specimens to better understand the molecular drivers of human disease. These proteomic technologies, including aptamer and immunoassay-based platforms, have begun to inform novel targets for diagnosis, prognosis and risk stratification, treatment monitoring, and therapeutic intervention in neurodegenerative disease (*8–11*). To date, these efforts have primarily focused on discovery of pathology-specific biomarkers and molecular pathways for discriminating symptomatic neurodegenerative disease patients from controls. By applying these sensitive molecular screening methods in cohorts with deep behavioral phenotyping and individuals that span the continuum of typical cognitive aging to dementia, we can identify in-vivo biological targets that underly the neuroprotective properties of lifestyle behaviors, such as PA.

The present study measured 7,288 plasma proteins using the aptamer-based SomaScan platform in 65 cognitively unimpaired older adults from the UCSF Brain Aging Network for Cognitive Health (BrANCH) who also completed 30 days of actigraphy monitoring for objective quantification of PA. Differential regression and weighted gene co-expression network analysis (WGCNA) identified communities of plasma proteins that were robustly associated with PA. Associations of plasma proteins with PA were validated by leveraging plasma proteomic datasets from two independent cohorts with different methods of PA quantification, an endurance exercise training intervention in young to middle aged adults (HERITAGE trial (*12*)) and an observational study of self-reported habitual PA in racially diverse middle to older aged adults (Atherosclerosis Risk In Communities [ARIC] study (*13*)). We next leveraged plasma proteomic, brain tissue proteomic, and genomic datasets from Alzheimer’s disease and related dementias (ADRD) cohorts to identify the molecular overlap between PA-related molecular pathways and ADRD risk. We identified individual plasma targets as well as broader communities of co-expressed proteins that robustly tracked with exercise, differed across symptomatic neurodegenerative conditions, prognosticated longitudinal cognitive decline, and correlated with brain tissue proteomic pathways linked to cognitive resilience. These PA proteomic signatures, which may underlie the neuroprotective effects of exercise on brain aging, were enriched for processes linked to cell adhesion, extracellular matrix (ECM), vascular remodeling, growth factor signaling, and immune response. Together, these data support the utility of leveraging exercise proteomics as a tool to probe biology relevant for dementia prevention.

## RESULTS

### UCSF BrANCH cohort characteristics

Table S1 reports demographic and clinical characteristics for the 65 cognitively unimpaired older adults (Clinical Dementia Rating = 0) completed Fitbit^TM^ monitoring and plasma proteomics through the UCSF BrANCH study. Participants were on average 76.7 years old (range = 59-91) with 17.6 years of education, 60% female, and a majority identified as non-Hispanic White (88%). Average daily step counts ranged from 669 to nearly 13,845 steps per day, with a sample mean of 7,277 steps per day.

### Network analysis highlights extracellular matrix and lipid homeostasis signatures of physical activity

We performed age- and sex-adjusted differential regressions to identify individual plasma proteins that were positively or negatively associated with PA (Table S2). Roughly 8% of the plasma proteome was nominally associated with PA (unadjusted *p* < .05), with 210 proteins positively associated with step count and 358 proteins negatively associated with step count (Fig. 1A-B). Six proteins survived FDR-correction, including higher abundance of anthrax toxin receptor cell adhesion molecules 1 and 2 (ANTXR1, ANTXR2; aka, capillary morphogenesis gene 1 and 2, respectively) as the top two PA hits by statistical significance. GO analysis of PA-associated proteins (Fig. 1C; Table S3) revealed strong links to lipoprotein and extracellular matrix (ECM) biology, including plasma lipoprotein particle clearance and cell adhesion (increased with PA), as well as lipoprotein particle receptor binding and ECM assembly (decreased with PA).

**Figure 1.**
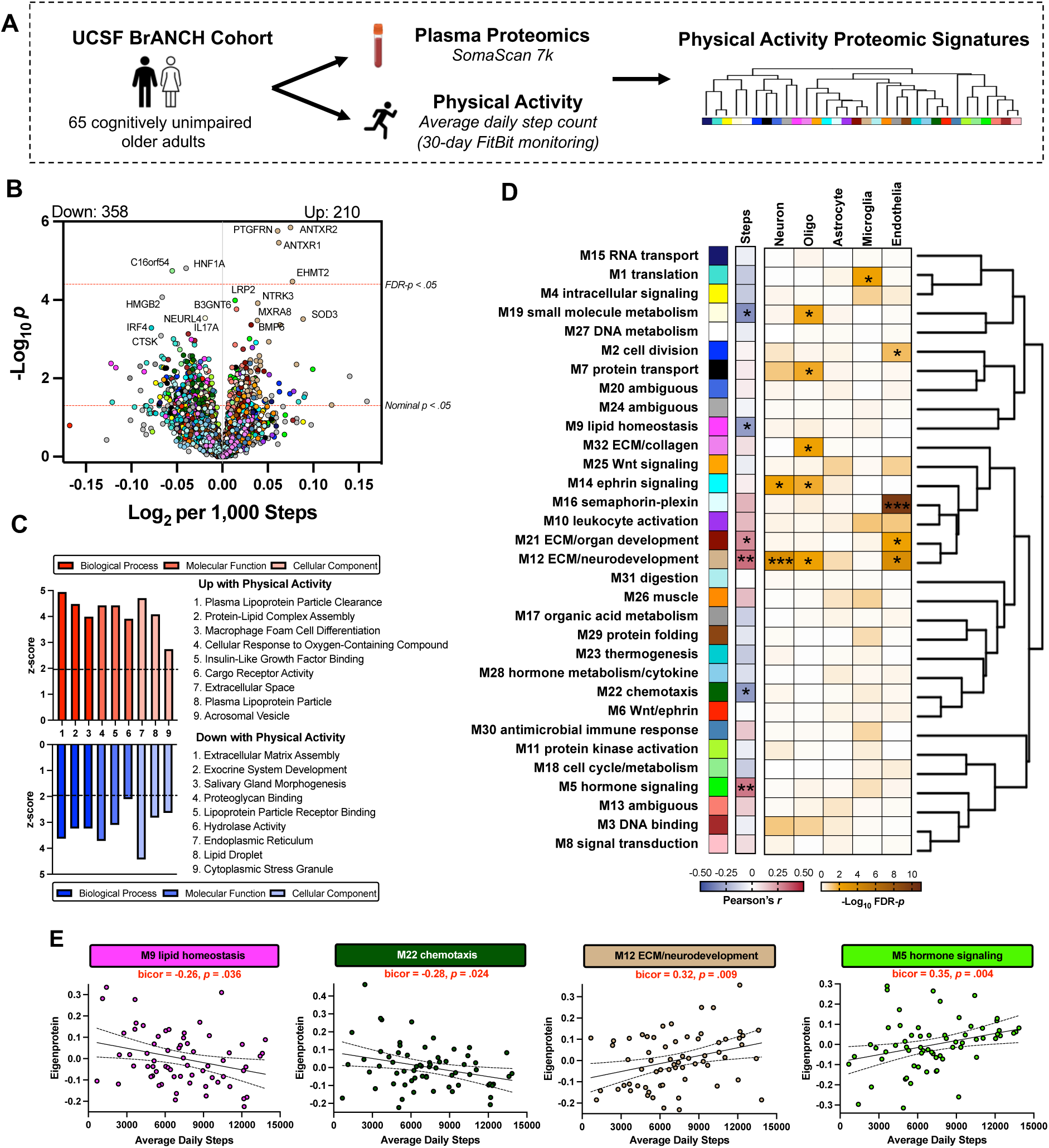
Plasma proteomic signatures of physical activity. **(A)** Plasma samples from 65 cognitively unimpaired adults in the UCSF Brain Aging Network for Cognitive Health (BrANCH) were analyzed using large-scale aptamer-based proteomics (SomaScan v4.1 [7k proteins]). Participants also completed 30-day Fitbit monitoring for quantification of physical activity, operationalized as average daily step count, which was integrated with proteomic data to identify plasma proteomic signatures of physical activity. (**B)** Differential regression models examined correlations between average daily step count and plasma protein abundance, adjusting for age and sex (see Table S2). Volcano plot depicts the log_2_-fold change in protein abundance for a 1,000 step increase in average daily step count by negative log_10_ *p*-value. Proteins are colored by the co-expression module they were assigned according to weighted gene co-expression network analysis (WGCNA). (**C)** Gene ontology (GO) analysis identified biological pathways associated with proteins that increased or decreased with higher physical activity (see Table S3). Enrichment for a given ontology is shown by *z* score, transformed from a Fisher’s exact test. (**D)** WGCNA identified 32 plasma protein co-expression modules (see Table S4). GO analysis was used to identify the principal biology represented by each module (see Table S5) and module relatedness is shown in the dendrogram to the right. Heatmaps represent module eigenprotein correlations with physical activity (see Table S6) and module-wise cell type enrichment, assessed by module protein overlap with cell-type-specific marker lists of neurons, oligodendrocytes, astrocytes, microglia and endothelia. \*\*\**p*<.001, \*\**p<*.01, \**p*<.05. (**E)** Scatterplots of physical activity correlated against select plasma protein co-expression modules.

We next leveraged WGCNA to identify communities of plasma proteins that were co-expressed across plasma samples and associated with PA. WGCNA revealed 32 protein co-expression modules (Fig. 1D). Protein module membership assignments are provided in Table S4. Pathway and cell type enrichment analyses were performed to identify the primary ontology used for module annotation (Table S5; Fig. S1). Nine modules exhibited significant overrepresentation for individual PA-associated proteins identified in differential regression (FDR-*p* < 0.05), and of those nine, six had module eigenproteins that were significantly correlated with PA. Module 12 (M12) ECM/neurodevelopment was comprised of 165 proteins and enriched for ontology terms related to neurodevelopment and ECM/cell adhesion, as well as neuronal, oligodendrocyte, and endothelial cell type markers. M12 module eigenprotein levels positively correlated with PA (bicor = 0.32, *p* = 0.009; Fig. 1E) and M12 also harbored the highest proportion of individual proteins that were positively associated with PA (54 out of 165 proteins [33%]), including four proteins that reached FDR significance from differential regression (ANTXR1, ANTXR2, prostaglandin F2 receptor inhibitor [PTGFRN], euchromatic histone-lysine N-methyltransferase 2 [EHMT2]). PA was also positively correlated with M21 ECM/organ development (bicor = 0.25, *p* = 0.046), enriched for endothelial cell types and closely related to M12, and M5 hormone signaling (bicor = 0.35, *p* = 0.004). M19 small molecule metabolism (bicor = −0.30, *p* = 0.015), M9 lipid homeostasis (bicor = −0.26, *p* = 0.036), and M22 chemotaxis (bicor = −0.28, *p* = 0.024) exhibited the strongest negative associations with PA.

### Physical activity proteomic signatures converge with age-related neurobehavior

We next correlated plasma proteins with neurobehavorial outcomes (cognitive composites, mood) and tested for module-wise overrepresentation of neurobehavioral-associated proteins in the BrANCH cohort of cognitively unimpaired older adults (Fig. 2A). All three modules that were positively associated with PA (M21, M12, M5) also harbored proteins that were positively associated with neurobehavior (Fig. 2B: executive function (M21, M12, M5), memory (M5), processing speed (M21, M12), and depressive symptoms (M12). To determine whether individual proteins linked to neurobehavior were also of high influence (‘hub proteins’) in M21, M12, and M5, we plotted individual protein correlational effect sizes against their intramodular connectivity; we focused on executive function given its relationship to PA (bicor = 0.35, *p*= 0.006) and all three modules. For all three modules, proteins with higher intramodular connectivity exhibited larger correlations with executive function (Fig. 2C), including M21 and M12 proteins linked to growth factor signaling/neurodevelopment (M21: NCAM1, BMP5, BMP6; M12: PRTG, ROBO1) and M5 proteins involved in immune response (CLEC4G, TNFRSF11A). Of the three modules that were negatively associated with PA, only M22 chemotaxis exhibited overrepresentation of proteins negatively correlated with neurobehavior, specifically memory and depressive symptoms. Taken together, these results demonstrate molecular overlap between PA and age-related neurobehavior, reflected by shared plasma proteomic signatures linked to neurodevelopment and immune signaling.

**Figure 2.**
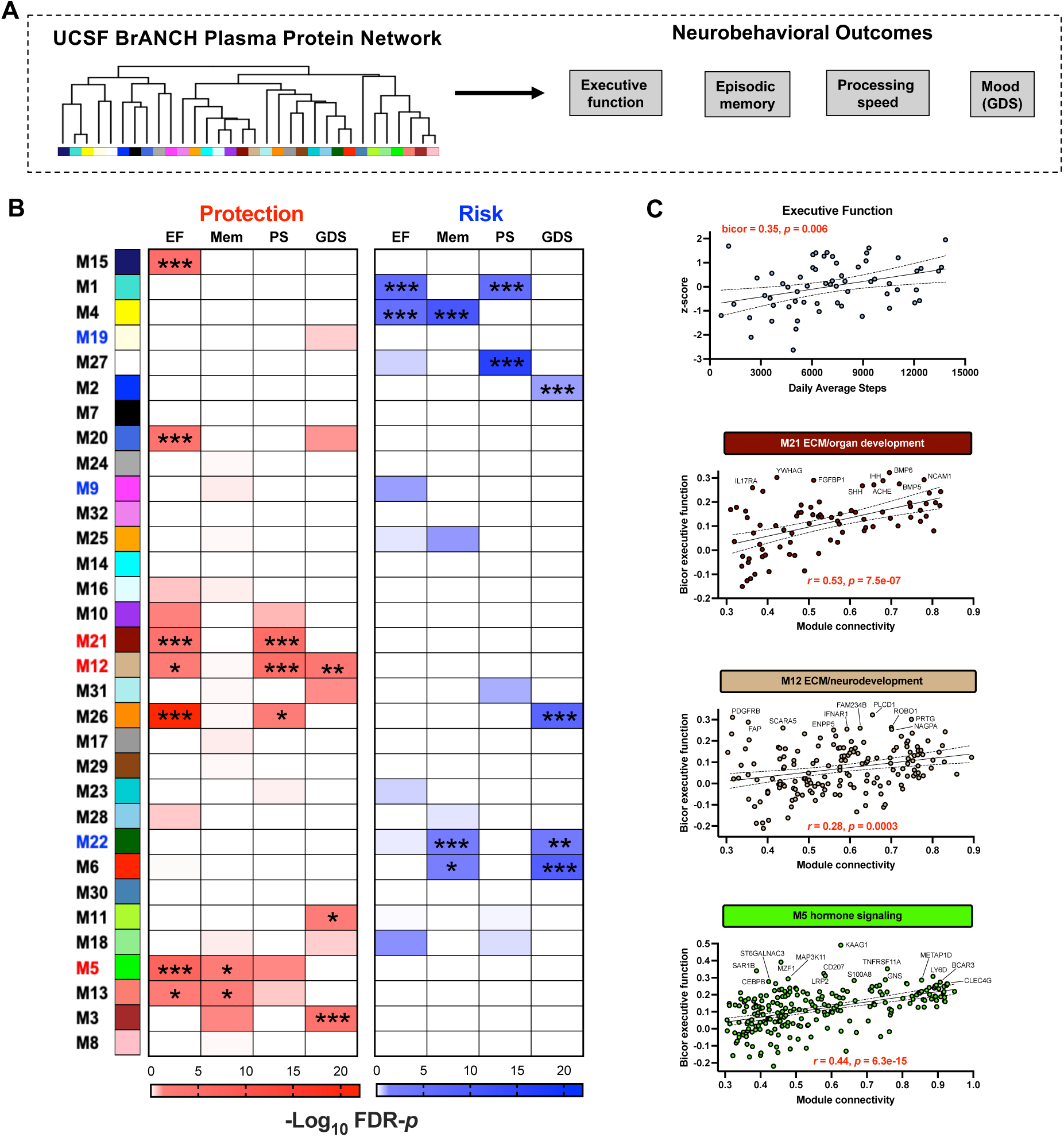
Physical activity-related plasma proteins predict age-related neurobehavior. **(A)** Plasma network module proteins were correlated against neurobehavioral outcomes measured in BrANCH (executive function, episodic memory, processing speed, mood [Geriatric Depression Scale]). (**B)** BrANCH plasma network module protein overlap with proteins positively (protection) or negatively (risk) correlated with neurobehavior. One-tailed Fisher’s exact test was used to determine module-wise enrichment, and results FDR-corrected using the Benjamini-Hochberg method (see Table S7). **(C)** Scatterplots display the correlation between physical activity and executive function as well as an individual protein’s strength of connectivity to a module (x-axis) against the individual protein’s correlation with executive function (y-axis). Annotated proteins are module members that were significantly correlated with executive function (bicor *p*<.05).

### Physical activity proteomic signatures replicate across cohorts and methods of physical activity quantification

The plasma proteomic signatures of PA identified in the discovery BrANCH cohort were based on observational actigraphy monitoring of PA in older adults. To determine whether these PA plasma proteomic signatures were robust to cohort composition and method of PA quantification, we cross-referenced summary statistics from two independent studies that examined plasma SomaScan proteins and PA (Fig. 3A): 1) the young to middle-aged HERITAGE exercise trial (N=654) with plasma proteins measured pre- and post-endurance training intervention(*12*), and 2) middle to old-aged ARIC cohort (N=10,644) with plasma proteins correlated against self-reported habitual PA(*13*). Most modules linked to PA in the Fitbit-based discovery BrANCH cohort were enriched for proteins linked to PA in the two independent datasets, with strong concordance in directionality (Fig. 3B). Specifically, M12 and M21 were enriched for proteins that increased post-intervention in HERITAGE and proteins positively correlated with self-reported PA in ARIC (FDR-*p* < 0.05). PA-associated proteins from HERITAGE and ARIC that overlap with M12 and M21 module proteins by intramodular connectivity are shown in Fig. 5C. M17 and M9 were enriched for proteins that decreased post-intervention in HERITAGE and, alongside M22, proteins negatively correlated with self-reported PA in ARIC (FDR-*p* < 0.05).

**Figure 3.**
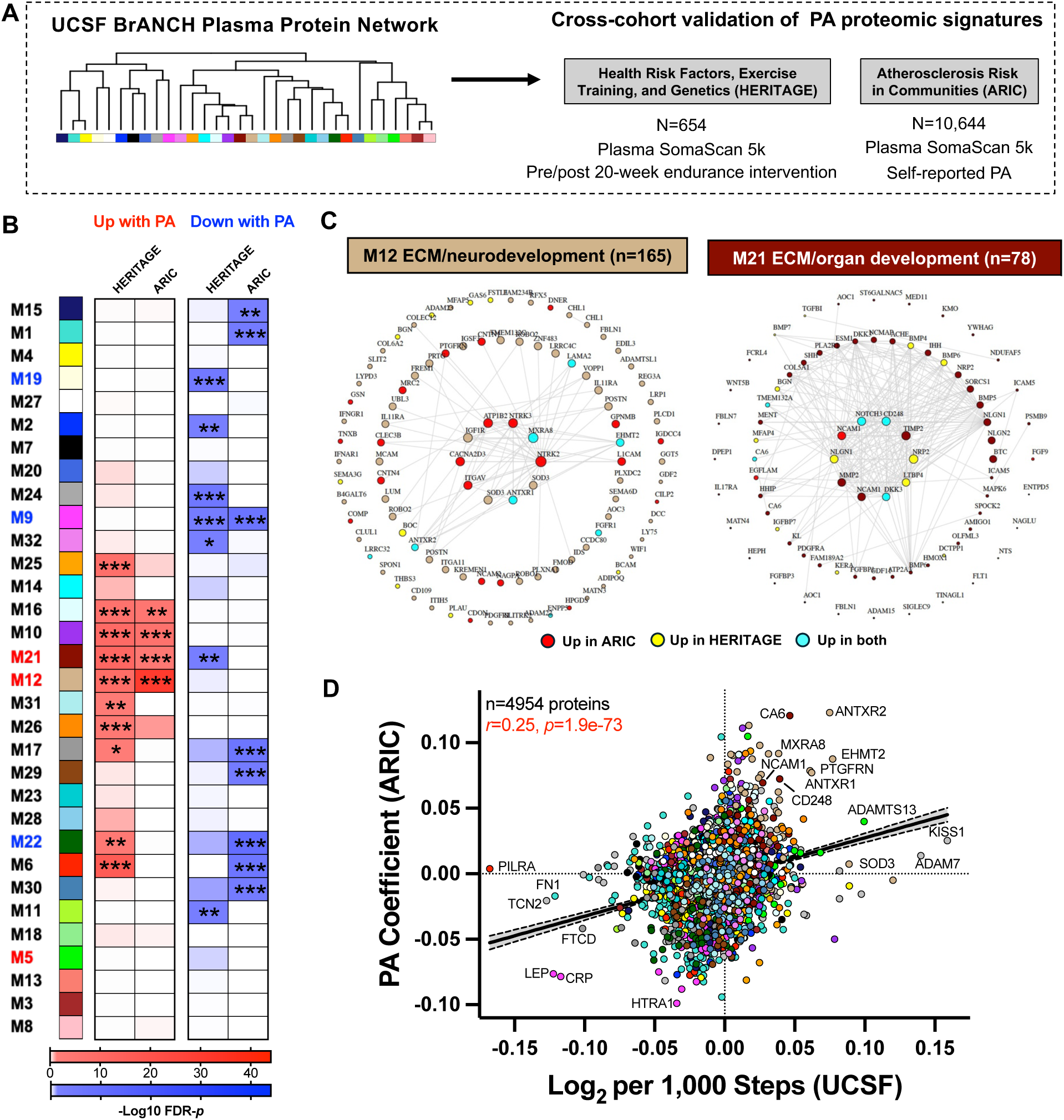
Physical activity plasma proteomic signatures are conserved across cohorts and methods of physical activity quantification. **(A)** BrANCH plasma network module proteins were cross-referenced with summary statistics from two independent studies that examined plasma proteomic (SomaScan v4.0 [5k]) correlates of an endurance training intervention (HERITAGE; Robbins et al. (*12*)) and self-reported physical activity (ARIC; Steffen et al. (*13*)). **(B)** BrANCH plasma network module protein overlap with proteins that changed pre/post HERITAGE intervention(*12*) or correlated with self-reported PA in ARIC(*13*). One-tailed Fisher’s exact test was used to determine module-wise enrichment, and results FDR-corrected using the Benjamini-Hochberg method (see Table S8). \*\*\**p*<.001, \*\**p<*.01, \**p*<.05. (**C)** Top 100 proteins by module connectivity (i.e., correlation with module eigenprotein value) for the M12 ECM/neurodevelopment and M21 ECM/organ develoment modules. Larger circles represent larger correlations, or module “hub” proteins. Proteins that increased in HERITAGE, ARIC, or both are highlighted. (**D)** Fitbit-based physical activity effect sizes in BrANCH were correlated against self-reported physical activity effect sizes in ARIC (N=4,954 overlapping protein measurements).

Correlation of Fitbit-based coefficients in BrANCH to self-reported PA-based coefficients in ARIC (N=4,954 overlapping protein measurements) also supported the concordant directionality of PA plasma proteomic signatures (*r* = 0.25, *p* = 1.9e-73; Fig. 3D). ANTXR2, the top hit in BrANCH, was also the top hit in ARIC in terms of statistical significance. In addition to ANTXR2, other hub proteins from M12 (matrix remodeling-associated protein 8 [MXRA8], EHMT2, ANTXR1, PTGFRN, neurotrophic tyrosine kinase receptor type 3 [NTRK3]) and M21 (CD248, neural cell adhesion molecule 1 [NCAM1]) exhibited strong associations with PA in ARIC.

### M12 ECM/neurodevelopment proteins are decreased across multiple neurodegenerative diseases

After establishing the cross-cohort reproducibility of PA plasma proteomic signatures and their convergence with neurobehavioral aging in cognitively unimpaired older adults, we next asked whether alterations in PA plasma proteomic signatures could be detected in symptomatic patients with Alzheimer’s disease and related dementias (ADRD). Thus, we leveraged plasma SomaScan datasets from the Stanford ADRC, which included patients with symptomatic AD and Parkinson’s disease (PD), and the ALLFTD Consortium, which included patients with symptomatic familial or sporadic frontotemporal lobar degeneration (FTLD) spectrum disorders (Fig. 4A). Module overrepresentation analysis indicated that M12 was enriched for plasma proteins with lower abundance across most ADRD groups compared to controls (FDR *p* < 0.05; Fig. 4B). Similarly, M21 exhibited a pattern of enrichment for proteins with lower abundance in *C9orf72*, *MAPT*, and sporadic progressive supranuclear palsy-Richardson’s syndrome (PSP-RS).

**Figure 4.**
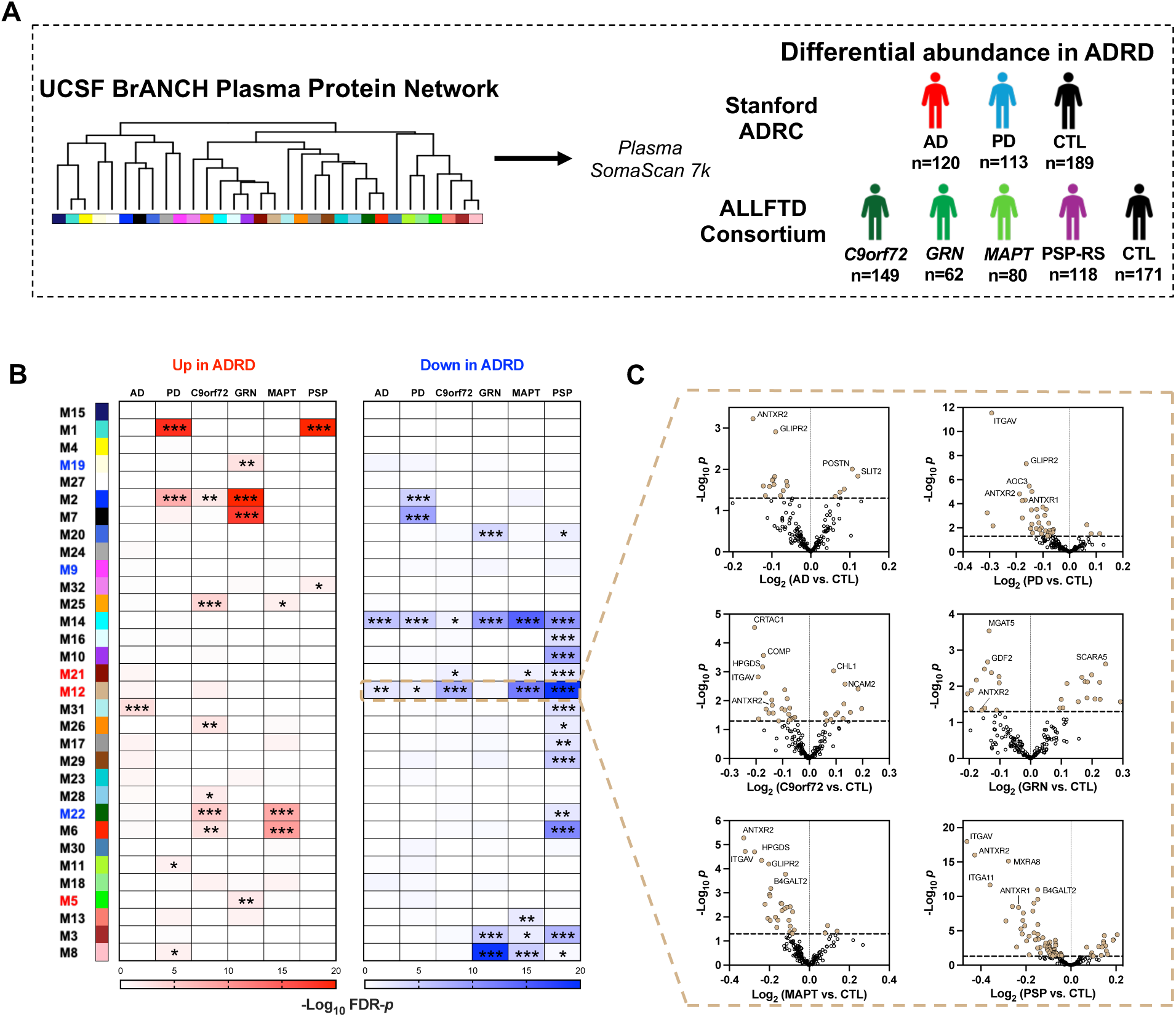
Physical activity plasma proteomic signatures are altered across Alzheimer’s disease and related dementias. **(A)** BrANCH plasma network module proteins were cross-referenced with differentially abundant plasma proteins in symptomatic Alzheimer’s disease and related dementias (ADRD). ADRD plasma proteomics data (SomaScan v4.1 [7k]) was sourced from two independent datasets, the Stanford ADRC (AD, Parkinson’s disease [PD], control) and the ALLFTD consortium (genetic [*C9orf72*, *GRN*, *MAPT*] and sporadic frontotemporal lobar degeneration [PSP-RS], controls). **(B)** BrANCH plasma network module protein overlap with proteins that increased or decreased in symptomatic ADRD. One-tailed Fisher’s exact test was used to determine module-wise enrichment, and results FDR-corrected using the Benjamini-Hochberg method (see Table S9). \*\*\**p*<.001, \*\**p<*.01, \**p*<.05. (**C)** Volcano plots display differential abundance of M12 ECM/neurodevelopment proteins in each ADRD group.

We next more deeply investigated individual proteins from M12, given the strong pattern of findings demonstrating its enrichment for plasma proteins that were reliably higher with PA in healthy adults and reliably lower across ADRD groups. First, we examined volcano plots of differential abundance of M12 proteins in each ADRD group (vs. controls) to identify individual proteins driving the observed patterns (Fig. 4C). ANTXR2 and integrin subunit alpha V (ITGAV), another M12 hub protein related to PA, both exhibited lower abundance across all six ADRD conditions compared to controls. ANTXR2 was the top M12 hit in AD and *MAPT* mutation carriers, and ITGAV was the top hit in PD and PSP-RS. In addition to ANTXR2 and ITGAV, seven other ECM and cell adhesion-related proteins from M12 that positively related to PA in prior analysis also exhibited lower abundance in over half of the ADRD groups examined (i.e., 4 or more): ANTXR1, ITGA11, MXRA8, GLIPR2, MGAT5, CLEC3B, and HPGDS.

### Plasma ANTXR2 predicts cognitive decline and AD brain proteomic signatures of cognitive resilience

We identified nine key M12 proteins that positively related to PA and were reliably lower in the symptomatic stage of multiple ADRDs; however, it remained unclear whether these plasma protein signals in symptomatic patients could predict future cognitive decline and reflected brain-based pathophysiological processes. To address these questions, we leveraged plasma proteomic (SomaScan) and brain tissue proteomic (TMT mass spectrometry(*14*)) data from 436 ROSMAP participants (baseline diagnoses: N=327 cognitively unimpaired, N=80 mild cognitive impairment, N=28 dementia) who completed antemortem blood draws and longitudinal cognitive assessments prior to brain donation (Fig. 5A). Cognitive analyses were first performed in the full sample and next in the subset of participants who were cognitively unimpaired at the time of blood draw, allowing us to test the prognostic value of PA-associated M12 hits measured during the presymptomatic phase of disease. Adjusting for ten neuropathologic indices, higher antemortem levels of ANTXR2, HPGDS, and CLEC3B significantly predicted slower global cognitive decline (Fig. 5B). In the subset of individuals who were cognitively unimpaired at baseline, only higher levels of ANTXR2 significantly predicted slower global cognitive decline (Fig. 5C).

**Figure 5.**
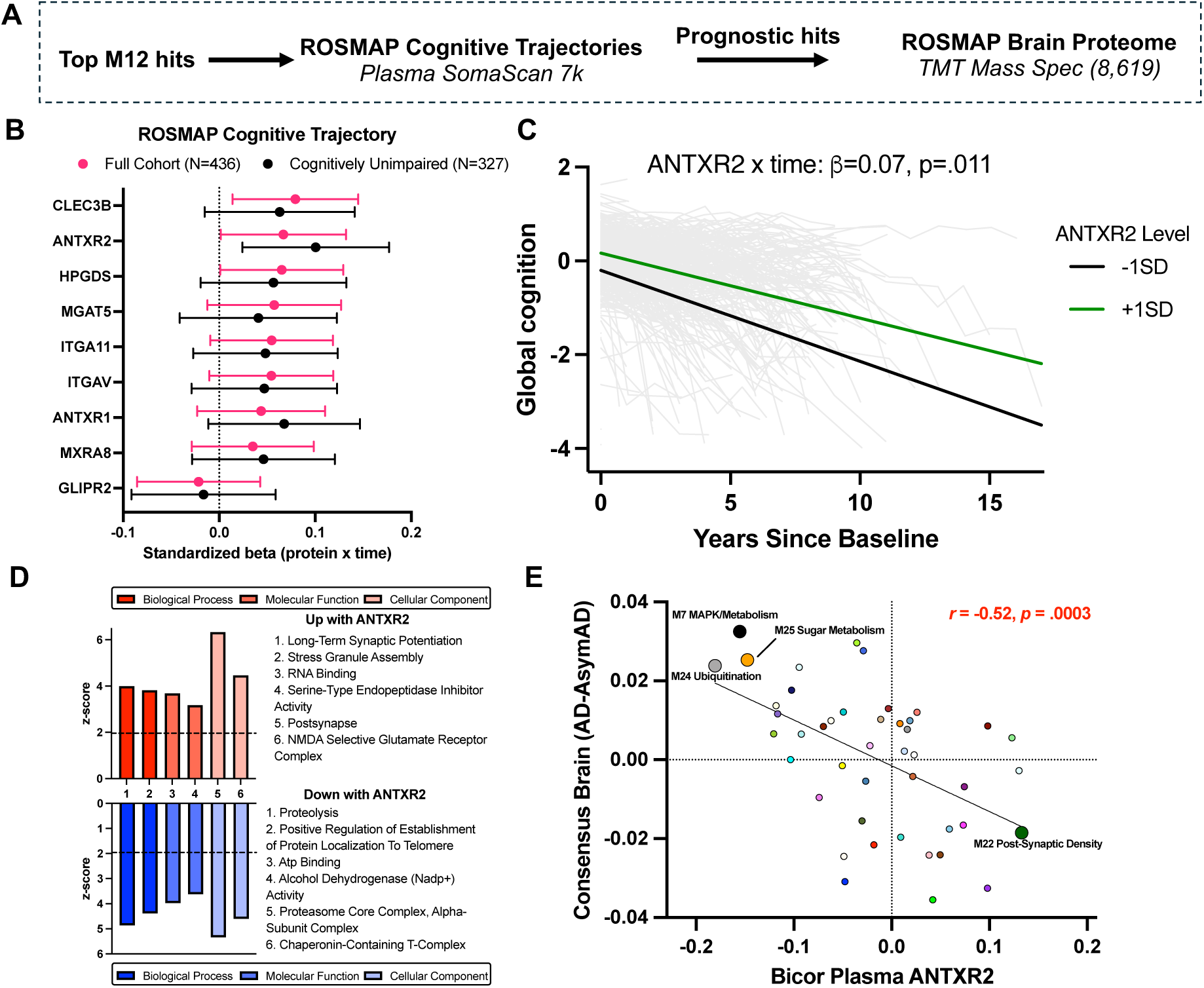
Plasma ANTXR2 predicts cognitive trajectories and brain tissue protein signatures in ROSMAP. (**A)** Plasma proteomics (SomaScan v4.1 [7k]) were measured at baseline in 436 ROSMAP participants who completed antemortem longitudinal cognitive testing and postmortem dorsolateral prefrontal cortex brain tissue proteomics (tandem mass tag mass spectrometry (TMT-MS) proteomics(*14*)). Top plasma hits from M12 were entered as predictors of global cognitive trajectories and brain tissue protein abundance. (**B)** Forest plot displays effect sizes of M12 proteins on global cognitive trajectories (standardized beta of protein x time) in the full cohort and the subset without cognitive impairment at baseline (N=327). Lower and upper bars represent 95% confidence intervals. (**C)** Spaghetti plot displays global cognitive trajectories at high (1SD) and low levels (−1SD) of plasma ANTXR2 across the full cohort. (**D)** Bar plots display results of GO analysis for postmortem brain tissue proteins associated with higher or lower antemortem plasma abundance of ANTXR2. Enrichment for a given ontology is shown by *z* score, transformed from a Fisher’s exact test. (**E)** Scatterplot displays the effect size of plasma ANTXR2 correlations with ROSMAP brain tissue network modules against the differential abundance of brain tissue modules in symptomatic vs. asymptomatic AD (as reported by Johnson et al.(*14*)). Brain tissue modules that differed between symptomatic and asymptomatic AD and significantly correlated with ANTXR2 (see Table S10) are annotated and displayed with larger circles.

We next leveraged the ROSMAP TMT-MS brain dataset (*14*) to identify brain proteomic pathways associated with peripheral abundance of ANTXR2. Differential correlation across 8,619 TMT-MS brain proteins revealed enrichment of synaptic signaling pathways with higher plasma abundance of ANTXR2 (e.g., “long-term synaptic potentiation”) and enrichment of proteolytic pathways with lower plasma abundance of ANTXR2 (e.g., “proteolysis”; Fig. 5D). Notably, plasma ANTXR2 did not correlate with brain TMT-MS measurement of ANTXR2 (bicor*<*0.01, *p*=.962). To further inform brain-based pathways of cognitive resilience that would be relevant for dementia prevention, we examined correlations of plasma ANTXR2 with brain protein co-expression modules in the subset of cases with autopsy-confirmed AD (107 symptomatic AD, 148 asymptomatic AD). ANTXR2 was significantly correlated with 5 previously identified brain protein co-expression modules(*14*). ANTXR2 positively correlated with brain M22 post-synaptic density, which had higher abundance in asymptomatic AD vs. symptomatic AD, and negatively correlated with brain M7 MAPK/metabolism, brain M24 Ubiquitination, and brain M25 Sugar Metabolism, which had lower abundance in asymptomatic AD vs. symptomatic AD (Fig. 5E).

Collectively, these results indicate that blood-based ANTXR2 can forecast preclinical cognitive decline and converge with synaptic and proteolytic brain proteomic signatures of cognitive resilience. These findings may reflect the unique contributions of peripherally-derived ANTXR2, our top discovery-based hit for PA, given the poor correlation of ANTXR2 between blood and brain.

### M12 ECM/neurodevelopment is enriched for AD genetic risk loci

As an environmental factor, PA could in part influence disease pathophysiology by interacting with molecular pathways that are also regulated by genetic risk factors. To interrogate the molecular overlap between PA and genetic dementia risk, we tested PA-related plasma modules for enrichment of GWAS hits using MAGMA. We specifically focused on AD GWAS targets given the large power of the extant AD GWAS literature (*15*), and thus higher confidence in AD GWAS hits, as well as our findings linking PA plasma signatures to the ROSMAP AD brain proteome. Four plasma modules were significantly enriched for AD GWAS targets (MAGMA [FDR] *p* < 0.05), including PA-related modules M12 ECM/neurodevelopment and M19 small molecule metabolism (Fig. 6). M12 featured 41 gene products from candidate AD genes, including top PA hits ANTXR1 and EHMT2. GO analysis of these 41 AD risk loci found in M12 revealed ontological enrichment for “heparin binding” proteins, specifically APOA5, CLEC3B, FSTL1, SLIT3, and SOD3. M19, which negatively correlated with PA, featured 21 gene products from candidate AD genes. The MAGMA enrichment of M19 was strongly driven by APOE (-log_10_ GWAS *p* = 72.7), also a heparin binding protein; of note, all 4 SOMAmer targets for APOE protein resided in M19. GO analysis did not reveal a clear ontological signal for these 21 AD risk loci in M19. Taken together, integration of AD GWAS data with our plasma protein network again showcased M12 as harboring multiple PA-associated targets with potential causative links to AD, possibly via heparin binding processes known to influence amyloid fibril formation (*16, 17*).

**Figure 6.**
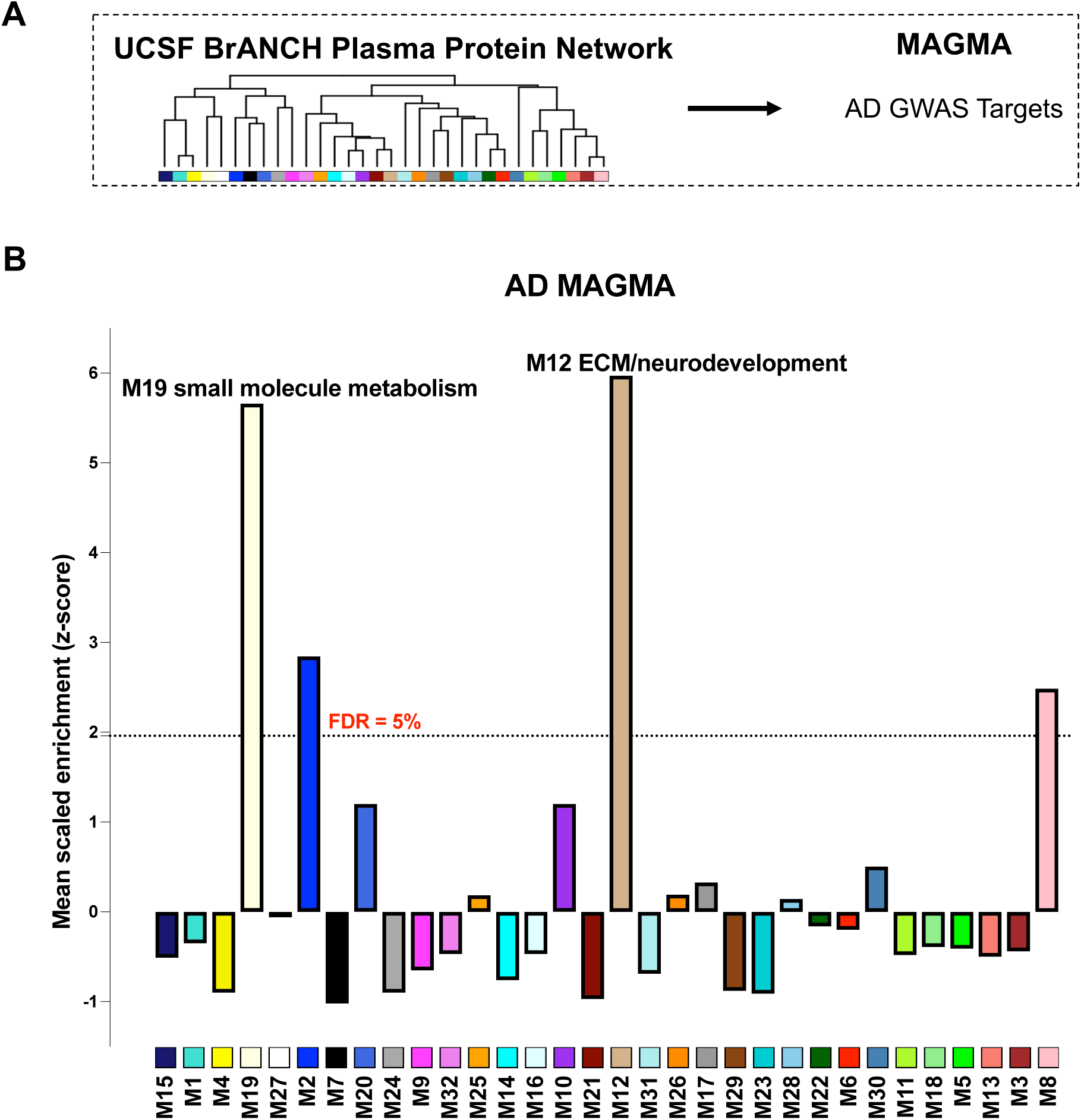
**(A)** BrANCH plasma network modules were tested for enrichment of AD genetic risk factor proteins using Multi-marker Analysis of GenoMic Annotation (MAGMA). Summary statistics were drawn from available AD genome wide association studies (GWAS)(*15, 51, 52*). (**B)** The dashed line indicates a mean scaled enrichment z-score of 1.96 (false discovery rate [FDR]-*p* = 0.05), above which enrichment was considered significant (see Table S10). Modules are ordered by relatedness as illustrated in Fig. 1D.

## DISCUSSION

This study integrated large-scale plasma proteomics with actigraphy-based step count data to identify molecular signatures of physical activity in aging adults. Our systems biology approach highlighted extracellular matrix (ECM) biology related to cell adhesion and angiogenesis, growth factor signaling, lipid homeostasis, and immuno-inflammatory biology as key pathways associated with objective physical activity levels. These pathways and individual proteins from PA-related co-expression modules were robustly associated with physical activity in two independent exercise cohorts, supporting the rigor and reproducibility of these exercise-related targets. In additional independent cohorts, the most concordant exercise-related proteins and pathways also differed by AD/ADRD diagnosis, predicted longitudinal cognitive trajectories and brain tissue proteomic signatures of cognitive resilience, and exhibited overlap with AD genetic risk loci, thereby highlighting a convergent biological link between blood-based markers of physical activity and neurodegenerative disease pathophysiology. ANTXR2, an extracellular matrix/cell adhesion molecule involved in angiogenesis and vascular remodeling (*18*), showed some of the strongest relationships with both physical activity and ADRD-related clinical and biological outcomes. Taken together, these findings 1) highlight molecules involved in ECM, lipid, and immunovascular pathways as potential exercise-associated AD/ADRD prevention targets, 2) advance the field towards biologically-informed implementation of lifestyle intervention for precision dementia prevention, and 3) integrates multiple bioinformatic techniques to develop a resource of exercise-related biology that warrants additional in-depth investigation.

The top exercise modules (M12 and M21) and individual proteins (e.g., ANTXR2/1, MXRA8, ITGAV) suggest a key role for activity-associated increases in peripheral ECM, particularly vascular remodeling, that associate with cognitive trajectories and are subsequently decreased across AD/ADRDs. These findings broadly align with the extant body of research linking physical activity to vascular health and further refine potential targets to changes in the structure and plasticity of the vascular matrix (*19*). Vascular ECM is comprised of a network of molecules that provide crucial structural and functional support for blood vessels, including maintaining integrity, elasticity, angiogenic repair/remodeling, and regulation of blood flow (*20*); a key target of exercise may therefore function through remodeling and rejuvenation of vascular molecules. These findings also align with an increasing number of proteomic studies in AD that are converging on both peripheral and brain ECM pathways as disease hubs. For instance, in autosomal dominant and sporadic AD, proteins in CSF and brain tissue reflecting ECM processes (e.g., SMOC1, SPON1) are consistently identified as top hits differentiating controls from AD, evidence dysregulation decades before symptom onset, and associate with early formation of amyloid plaques (*14, 21, 22*). Using unbiased plasma proteomics, our work further extends these findings to suggest a role for the *peripheral* ECM for brain health and highlights the ECM as a potential therapy target underlying how exercise may support dementia prevention in humans.

Further, within the M12 ECM/neurodevelopment module, ANTXR2, MXRA8, and ITGAV were among the top individual hits most strongly increased with physical activity and decreased across ADRD syndromes. ANTXR2, aka capillary morphogenic gene 2 (CMG2), was canonically discovered as a receptor for the anthrax toxin virus but was subsequently found to have key roles in normal physiology, including repair and stabilization of blood vessels, ECM organization and cell adhesion to ECM, and vascular tissue remodeling (*23, 24*). Similarly, MXRA8 is a cell surface protein also originally identified for its role as a receptor facilitating entry of arthritogenic alphaviruses (*25*). MXRA8 is strongly expressed in joint and skeletal muscle tissue and involved in response to tissue injury (*26*), raising the possibility that MXRA8 possesses “myokine” like properties with release from skeletal/muscle tissue following exercise related contractions. Integrins are heterodimeric membrane proteins composed of alpha and beta subunits that facilitate cell-ECM interactions. In the present study, the aptamer for ITGAV that demonstrate the strongest associations with PA and ADRDs targeted the ITGAV:ITGB3 heterodimer (Somalogic sequence id: 20187-10), which functions as a receptor for numerous ECM interacting proteins involved in angiogenesis (e.g., fibronectin) and hemostasis (e.g., vitronectin) (*27*). Together, these targets support the notion that exercise supports tissue related plasticity that associate with neuroprotective outcomes. It is unclear if these proteins cross the blood brain barrier and in fact, we did not observe a strong association between peripheral and brain tissue levels of ANTXR2; however, ANTXR2 did show significant associations with brain tissue proteins reflecting synaptic signaling and proteolytic functions, suggesting downstream neuroprotective associations in the CNS. Nonetheless, given the primarily observational nature of our data, future work is needed to determine the tissue and cellular origin and mechanistic importance of promising exercise molecules in non-human AD/ADRD models.

Other top exercise pathways that overlapped with brain aging outcomes in our study also highlighted molecules involved in bone and growth factor signaling (e.g., BMP5, BMP6, CLEC3B) as well as immune response (e.g., CLEC4G, TNFRSF11A). Immune benefits of physical activity are extensively documented (*28*). These data suggest additional specificity such that bone-associated immune signaling may link physical activity and cognitive outcomes. Indeed, emerging data support a bone-brain axis in which communication between bone-derived molecules may drive brain outcomes (*29*). In the CNS, physical activity is strongly linked to decreased microglial and astrocytic activation in animal models (*30–32*), and emerging data support the relevance of these exercise immune pathways in humans (*33*). Deeper investigation of how the immune system may serve as a communication axis between peripheral organ systems, including the bone, to brain following exercise are likely high yield avenues of future work.

Regarding study limitations, we exclusively leveraged SomaScan datasets for plasma proteomic analyses to ensure cross-cohort findings were not confounded by methodological differences; however, this approach limited our blood-based biological discovery and validation to targets measured on SomaScan, which may introduce bias in the proteomic pathways identified. Nonetheless, we showed that candidate plasma SomaScan hits in ROSMAP meaningfully associated with brain tissue proteomics assayed via an orthogonal technique, TMT-MS, supporting the potential generalizability and robustness of our findings. Another limitation is that our discovery cohort primarily consisted of highly educated white individuals. Despite this, our results were validated and showed consistency across a more diverse aging cohort using the ARIC Study. Importantly, with the exception of the HERITAGE exercise trial, most of the data incorporated in this study were observational. The directionality of the top exercise-pathways and individual targets on ADRD-related outcomes therefore cannot be currently determined, though our inclusion of genetic and longitudinal cognitive data helps mitigate this concern. Future research with mechanistic models is needed to explore the potential of candidate exercise markers as therapeutic targets for dementia prevention.

Identifying the biology of well-known resilience behaviors such as physical activity can be used to unlock new insights into dementia prevention and treatment. Here we provide an in-depth interrogation of peripheral exercise signatures for future studies to build upon. Further examination and experimental modulation of the identified proteins, including those represented in M12 ECM/neurodevelopment, are next steps towards validating these as potential biomarker and therapeutic targets. With an eye towards precision medicine, our findings also support the use of plasma proteomics to characterize individual biological signatures and identify people at greatest risk of adverse brain aging who may benefit most from exercise intervention.

## MATERIALS AND METHODS

### BrANCH Discovery Cohort Participants

Participants included 65 clinically normal, community-dwelling older adults enrolled in the UCSF Brain Aging Network for Cognitive Health (BrANCH) study. All participants underwent clinical evaluations including comprehensive history, neurologic exam, neuropsychological testing, caregiver interview and functional assessment (CDR) and blood draw. All participants were free of cognitive symptoms, lacked major neurological, psychiatric, or medical conditions (e.g., sleep apnea, stroke, HIV), were functionally intact (CDR=0), and were diagnosed as clinically normal per multidisciplinary consensus case conference (*34*). Study procedures were approved by the UCSF Committee on Human Research and all participants provided written informed consent.

### Plasma aptamer-based proteomics

Venous blood was collected and stored in EDTA tubes (Alzheimer’s Disease Neuroimaging Initiative protocol) at − 80 °C at UCSF until being packed with dry ice and sent to SomaLogic (SomaLogic, Boulder, CO) for proteomics analysis. Plasma proteins were measured on the SomaScan v4.1 platform (*35*), which included 7,288 targets across 6,358 unique proteins. The SomaScan assay leverages slow off-rate modified aptamers (SOMAmers), which are short, single stranded deoxynucleotides that bind to protein targets with high specificity (*36*). A volume of 65 microliters of plasma was used to create SOMAmer – protein reactions in 96-well plates. Tagged SOMAmer-protein complexes were captured in a bead-based assay, and levels of SOMAmer bound to sample were quantified through a fluorescent signal in DNA hybridization microarrays (*35*). The reaction signal was detected digitally and expressed as aggregated Agilent relative fluorescent units (RFU), which were normalized to scale and subsequently log_2_-transformed.

Quality control was performed using a previously published pipeline that identified protein values below the target-specific limit of detection (LOD) (*8*), which was defined as median log_2_ buffer signal plus 3 standard deviations (SD) of the assay’s buffer measurements. SomaScan background SD was calculated from available buffer replicate data for each target. Consistent with prior work demonstrating optimization of the SomaScan platform for plasma samples (*8*), all targets had low rates of values below LOD and thus all 7,288 targets were retained for downstream analysis.

### Physical activity quantification

Participants were asked to complete 30 continuous days of observational Fitbit™ Flex 2 PA monitoring during waking hours. Fitbit data were quality-checked and cleaned, which included controlling for non-adherence by excluding days when there was a high suspicion that participants did not wear the device per previously published methods (i.e., days with <100 steps total) (*37, 38*). Only participants who were adherent to at least 14 days of Fitbit monitoring were included. Total step counts per day collected from the Fitbit device were averaged to quantify the average daily step count.

### Differential regression analysis

General linear models examined the relationship between average daily steps and plasma abundance for each protein separately, adjusting for age and sex. Model coefficients were multiplied by 1,000 such that values represent the log_2_-fold protein increase/decrease associated with an increase/decrease of 1,000 average daily steps. PA-related proteins were identified based on nominal statistical significance (*p* < 0.05) and top hits were further identified based on Benajmini-Hochberg false discovery rate (FDR) corrected *p* < 0.05.

### Plasma protein co-expression network analysis

WGCNA(*14, 39*) was used to generate a plasma protein network from the *n*=7,288 log_2_ protein abundance × *n*=65 case samples matrix. No outliers were detected using the WGCNA sample network connectivity outlier algorithm (*14*). The WGCNA blockwiseModules function was run with the following parameters: power=12, deepSplit=4, minModuleSize=10, mergeCutHeight=0.07, TOMdenom=“mean”, bicor correlation, signed network type, PAM staging and PAM respects dendro as TRUE, with clustering completed within a single block. Module memberships were then iteratively reassigned to enforce kME table consistency, as previously described (*14*).

### Gene ontology (GO) and cell type marker enrichment

GO enrichment of differential abundance lists and network modules was calculated as a Fisher exact test p value transformed to z score and visualized using the publicly-available GOparallel code (https://www.github.com/edammer/GOparallel), which pulls curated gene sets from the Bader Lab’s monthly update (*40*). As previously published (*14*), enrichment of network modules for cell type-specific marker gene symbols was performed using one-tailed Fisher’s exact test (https://github.com/edammer/CellTypeFET).

### BrANCH neurobehavioral assessment

BrANCH participants underwent detailed neurobehavioral assessments, including comprehensive neuropsychological testing and self-report questionnaires. Cognitive performance was operationalized using sample-based z-score composites of episodic memory, executive functioning, and processing speed, as previously described (*41*). Cognitive composites were regressed for age, sex, and education based on the full sample of clinically normal BrANCH participants (N > 488 per composite), and residualized z-scores were examined as cognitive outcomes in the present study. Processing speed z-scores were reverse coded such that higher scores translated to better (faster) performance, consistent with other cognitive outcomes. Participants also completed the 30-item self-report Geriatric Depression Scale (GDS), which quantified mood symptoms (*42, 43*). GDS total scores were also regressed for demographics and reverse coded for analysis such that higher scores translated to better (less symptomatic) mood. The *bicorAndPvalue* function from the *R* WGCNA package was used to calculate biweight midcorrelations (bicor) between average daily steps and neurobehavioral outcomes, as well as individual plasma proteins and each neurobehavioral outcome. One-tailed Fisher’s exact tests determined plasma network module-wise overrepresentation of proteins positively (protection) or negatively (risk) associated with neurobehavioral outcomes, adjusting for FDR.

### Validation of physical activity proteomic signatures in HERITAGE and ARIC

To evaluate the cross-cohort validity of our PA plasma proteomic signatures, we compared our findings to summary statistics from two complementary studies examining SomaScan plasma proteomics and PA. The first study by Robbins et al. examined plasma protein changes before and after a 20-week endurance exercise training intervention in 654 healthy adults from the Health Risk Factors, Exercise Training, and Genetics (HERITAGE) cohort (ages 17-65 years) (*12*). The second study by Steffen et al. examined plasma proteomic correlates of self-reported PA, quantified via the Baecke Physical Activity Questionnaire, in 10,644 participants from the visit 3 examination of the Atherosclerosis Risk in Communities (ARIC) Study cohort (mean [SD] age: 60 [5.7] years)(*13*). Both studies included plasma proteomic data from the SomaScan v4.0 platform, which measured roughly 5,000 targets. One-tailed Fisher’s exact tests determined BrANCH plasma network module-wise overrepresentation of proteins positively or negatively associated with PA based on the HERITAGE intervention or ARIC self-reported PA. Since multivariable regression coefficients were available for all proteins measured in the ARIC study, we also correlated objectively monitored PA effect sizes (UCSF BrANCH) with self-reported PA effect sizes (ARIC) for the 4,954 proteins that overlapped across BrANCH and ARIC datasets.

### PA proteomic overlap in symptomatic disease cohorts: Stanford ADRC and ALLFTD Consortium

We leveraged plasma proteomics data (SomaScan v4.1) from two cohorts of patients with symptomatic neurodegenerative diseases to test whether PA-related plasma protein changes in the healthy aging BrANCH cohort overlapped with plasma protein signatures of ADRD: 1) The Stanford ADRC plasma SomaScan dataset included 120 patients with MCI or dementia due to AD, 113 patients with Parkinson’s disease (PD) and related disorders (i.e., Lewy body dementia), and 189 AD biomarker-negative controls; 2) The ARTFL/LEFFTDS Longitudinal Frontotemporal Lobar Degeneration (ALLFTD) consortium (NCT04363684) (*44, 45*) plasma SomaScan dataset included symptomatic carriers of pathogenic mutations in frontotemporal lobar degeneration (FTLD)-tau (*MAPT* [n=80]) or FTLD-TDP causing genes (*C9orf72* [n=149], *GRN* [n=62]), patients with sporadic progressive supranuclear palsy-Richardson’s syndrome (PSP-RS; n=118), which is highly specific for underlying PSP pathology (*46*), and noncarrier controls (n=171) from families with a known mutation in one of the FTLD-causing genes (*47*). Differential abundance analyses examined ADRD group differences in plasma protein abundance compared to controls. One-tailed Fisher’s exact tests examined overrepresentation of ADRD-associated proteins across BrANCH plasma network modules.

### Cognitive trajectory and brain proteome association analyses in ROSMAP

We leveraged antemortem plasma proteomic (SomaScan v4.1), longitudinal cognitive data, and postmortem brain tissue proteomic data from 436 participants in the Religious Orders Study and Rush Memory and Aging Project (ROSMAP) cohort to determine the association of PA-associated plasma proteins with cognitive trajectories and brain protein signatures of cognitive resilience. ROSMAP participants were without known dementia at baseline study entry and all agreed to detailed clinical evaluation and brain donation at death (*48*). Both studies were approved by an Institutional Review Board of Rush University Medical Center. Each participant signed an informed consent, Anatomic Gift Act, and an RADC Repository consent (allowing their data and biospecimens to be repurposed). Participant cognitive status at the blood draw ROSMAP timepoints used in the present analysis included 327 individuals without cognitive impairment, 81 with mild cognitive impairment, and 28 with dementia. Participants completed a median [IQR] of 5 [3 – 8] cognitive assessments spanning a median [IQR] of 4 [2 – 7] years of follow-up. All participants underwent autopsy with postmortem dorsolateral prefrontal cortex brain tissue assayed for 8,619 proteins using tandem mass tag mass spectrometry (TMT-MS) proteomics (*14*). Linear mixed-effects models examined the relationship between baseline plasma protein concentrations and longitudinal global cognitive trajectories (interaction of protein x time [years since plasma baseline]), adjusting for ten standard neuropathological indices measured in ROSMAP: amyloid-β, tangles, cerebral amyloid angiopathy, cerebral atherosclerosis, arteriolosclerosis, Lewy body, TDP-43, gross infarct, microinfarct and hippocampal sclerosis (*14, 49*). We next performed differential correlations between plasma protein levels and the 8,619 brain proteins across all 436 donors, adjusting for FDR. GO enrichment of brain proteomic correlates of plasma proteins was performed as described above. To determine whether cognitive trajectory-associated plasma protein levels converged with brain proteomic signatures of cognitive resilience, we examined plasma protein correlations with brain tissue protein co-expression modules (*14*) among the subset of donors with pathologically-confirmed AD (n=255), which included 107 brain donors who were symptomatic at death (AD) as well as 148 cognitively resilient donors who were asymptomatic at death (AsymAD). Results were visualized by plotting plasma protein correlation coefficients with brain tissue modules against AD vs. AsymAD brain module eigenprotein differences.

### AD GWAS module association

We determined whether plasma network modules linked to PA were also enriched for gene products of AD GWAS targets using MAGMA (version 1.09b) (*50*) and single nucleotide polymorphism (SNP) summary statistics from available AD GWAS studies (*15, 51, 52*), as previously described (*14, 53*). MAGMA was performed using publicly-available code: https://github.com/edammer/MAGMA.SPA. GWAS lists were filtered for genes with AD association values of *p* < 0.05 prior to MAGMA.

### Statistical Analysis

Statistical analyses were performed in R (v4.3.1). Correlations were performed using biweight midcorrelations (bicor) or Pearson’s correlations where indicated. Comparisons between two groups were performed by a two-sided t test. Comparisons among three or more groups were performed with ANOVA with Tukey’s pairwise comparison of significance. *P* values were adjusted for multiple comparisons by FDR correction according to the Benjamini-Hochberg method where indicated.

## Supporting information

Suppmentary Tables

## Data Availability

UCSF BrANCH data are available on reasonable request made to the UCSF Memory and Aging Center. Academic, not-for-profit investigators can request data for professional education and for research studies. Requests can be made online (https://memory.ucsf.edu/research-trials/professional/open-science). Datasets used for the analyses for the current study are also available from the corresponding author on reasonable request. Algorithms used for protein data processing and analysis are available in existing R packages and at https://github.com/edammer, as described in the Methods. ROSMAP resources can be requested at https://www.radc.rush.edu and www.synpase.org. Pre-existing data access policies for each of the ARIC parent cohort studies specify that research data requests can be submitted to each steering committee; these will be promptly reviewed for confidentiality or intellectual property restrictions and will not unreasonably be refused. Please refer to the data sharing policies of these studies. Individual level patient or protein data may further be restricted by consent, confidentiality or privacy laws/considerations. These policies apply to both clinical and proteomic data.

## Acknowledgments

We are deeply grateful to study participants and their families for participating in the UCSF Memory and Aging Center and other research programs from which data were sourced for this project. We additionally thank the teams of coordinators and administrative staffs who have provided integral support for the ongoing execution of research at our centers.

## Funding

National Institutes of Health grant R01AG032289 (JHK)

National Institutes of Health grant R01AG048234 (JHK)

National Institutes of Health grant P30AG062422 (GDR)

National Institutes of Health grant R01AG072475 (KBC)

National Institutes of Health grant UF1NS100608 (JHK)

National Institutes of Health grant K23AG058752 (KBC)

Alzheimer’s Association Research Grant AARG-20-683875 (KBC)

Larry L. Hillblom Foundation grant 2024-A-001-CTR (KBC)

Larry L. Hillblom Foundation grant 2018-A-006-NET (JHK)

Alzheimer’s Association grant AARF-23-1145318 (RS)

New Vision Research grant CCAD 2024-001-1 (RS)

American Academy of Neurology, Association for Frontotemporal Lobar Degeneration, and American Brain Foundation Fellowship Award (RS)

Alzheimer’s Association grant AARF-22-974065 (EWP)

National Institutes of Health grant K23AG084883 (EWP)

National Institutes of Health grant P30AG062422 (EWP)

Shenandoah Foundation (EWP)

ALLFTD Consortium grant U19AG063911 (funded by the National Institute on Aging and the National Institute of Neurological Diseases and Stroke)

ARTFL Consortium grant U54NS092089 (funded by the National Institute of Neurological Diseases and Stroke and National Center for Advancing Translational Sciences)

LEFFTDS Consortium grant U01AG045390 (funded by the National Institute on Aging and the National Institute of Neurological Diseases and Stroke)

National Institutes of Health grant P50AG047366 (Stanford ADRC)

National Institutes of Health grant P30AG066515 (Stanford ADRC)

National Institutes of Health grant P30AG10161 (ROSMAP)

National Institutes of Health grant P30AG72975 (ROSMAP)

National Institutes of Health grant R01AG15819 (ROSMAP)

National Institutes of Health grant R01AG17917 (ROSMAP)

National Institutes of Health grant U01AG46152 (ROSMAP)

National Institutes of Health grant U01AG61356 (ROSMAP)

Gates Ventures (ROSMAP)

National Heart, Lung, and Blood Institute, National Institutes of Health, Department of Health and Human Services, under Contract nos. 75N92022D00001, 75N92022D00002, 75N92022D00003, 75N92022D00004, 75N92022D00005 (ARIC study)

National Institutes of Health/National Heart, Lung, and Blood Institute grant R01HL134320

SomaLogic Inc. conducted the SomaScan assays in exchange for use of ARIC data. The funding sources had no role in the design and conduct of the study; in the collection, analysis, or interpretation of the data; or in the preparation, review, or approval of the manuscript.

## Author contributions

Conceptualization: RS, EWP, KBC

Methodology: RS, EWP, KBC

Investigation: RS, EWP, KBC

Visualization: RS

Funding acquisition: BFB, HJR, ALB, JHK, KBC

Writing—original draft: RS, EWP, AVB, CJC, CC, KBC

Writing—review & editing: RS, KBC

All authors contributed to acquisition, analysis, or interpretation of data or revision of the manuscript.

## Supplementary Materials

**Figure S1. Pathway analysis on BrANCH plasma network modules.** Gene ontology (GO) analysis was performed to ascertain the principal biology represented by the constituent proteins in each module. Enrichment for a given ontology is shown by z score, transformed from a Fisher’s exact test.

**Table S1.** BrANCH Cohort Characteristics

**Table S2.** Differential Regression of Physical Activity and the Plasma Proteome

**Table S3.** Differential Regression GO Term Enrichment

**Table S4.** Plasma Network Module Membership

**Table S5.** Plasma Network Module GO Term Enrichment

**Table S6.** Plasma Network Module Associations with Physical Activity

**Table S7.** Hypergeometric Overlap Statistics for Enrichment of Neurobehavioral Proteins in Plasma Network Modules

**Table S8.** Hypergeometric Overlap Statistics for Enrichment of Exercise Intervention and Self-Reported Physical Activity Proteins in Plasma Network Modules

**Table S9.** Hypergeometric Overlap Statistics for Enrichment of ADRD Plasma Proteins in Plasma Network Modules

**Table S10.** ROSMAP Plasma ANTXR2 Associations with TMT-MS Brain Proteome

**Table S11.** Alzheimer’s disease (AD) GWAS MAGMA

