## Supplementary figures and images for "Network-based Plasma Proteomics Reveals Molecular Overlap Between Physical Activity and Dementia Risk"

### Supplementary Figure 1

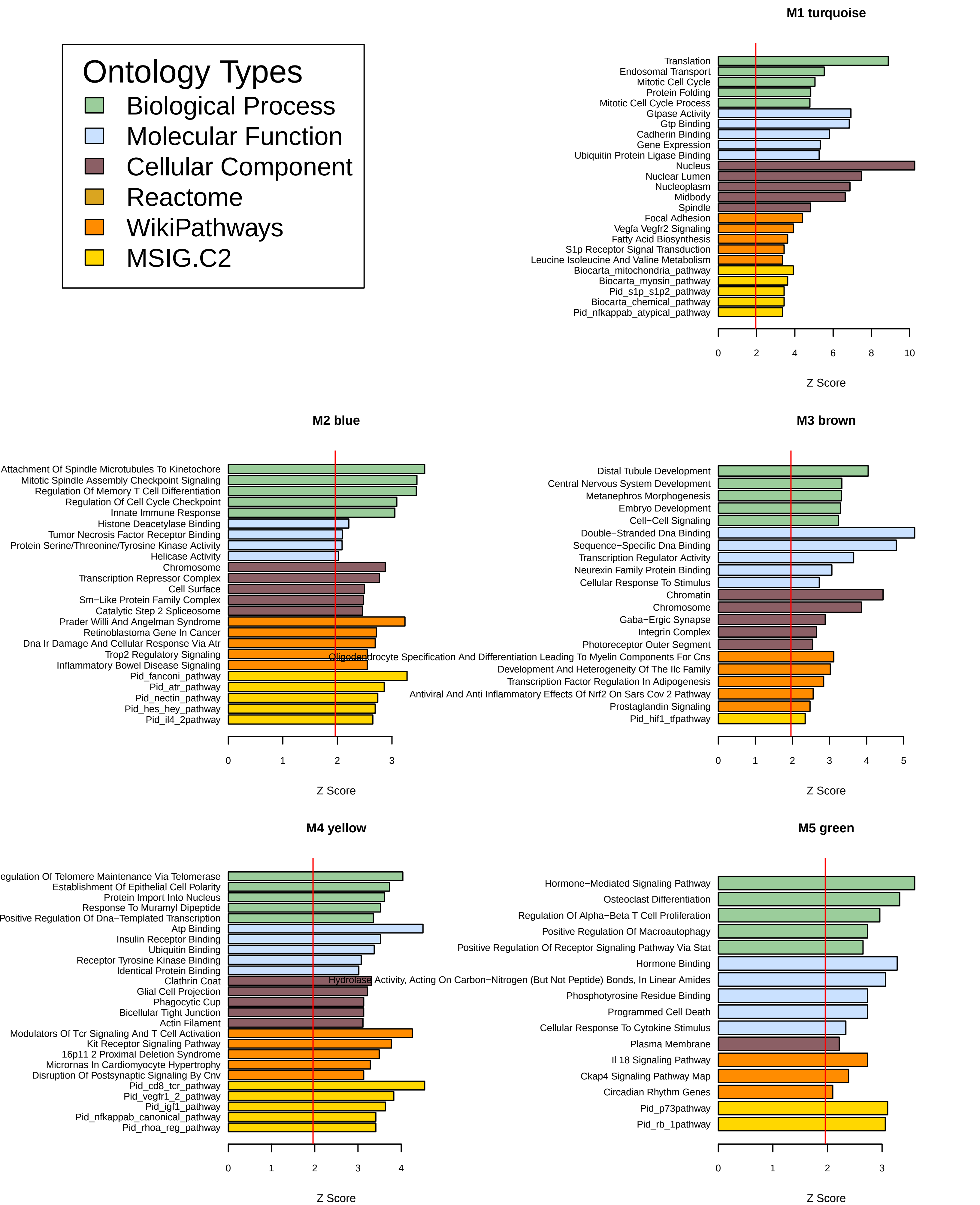

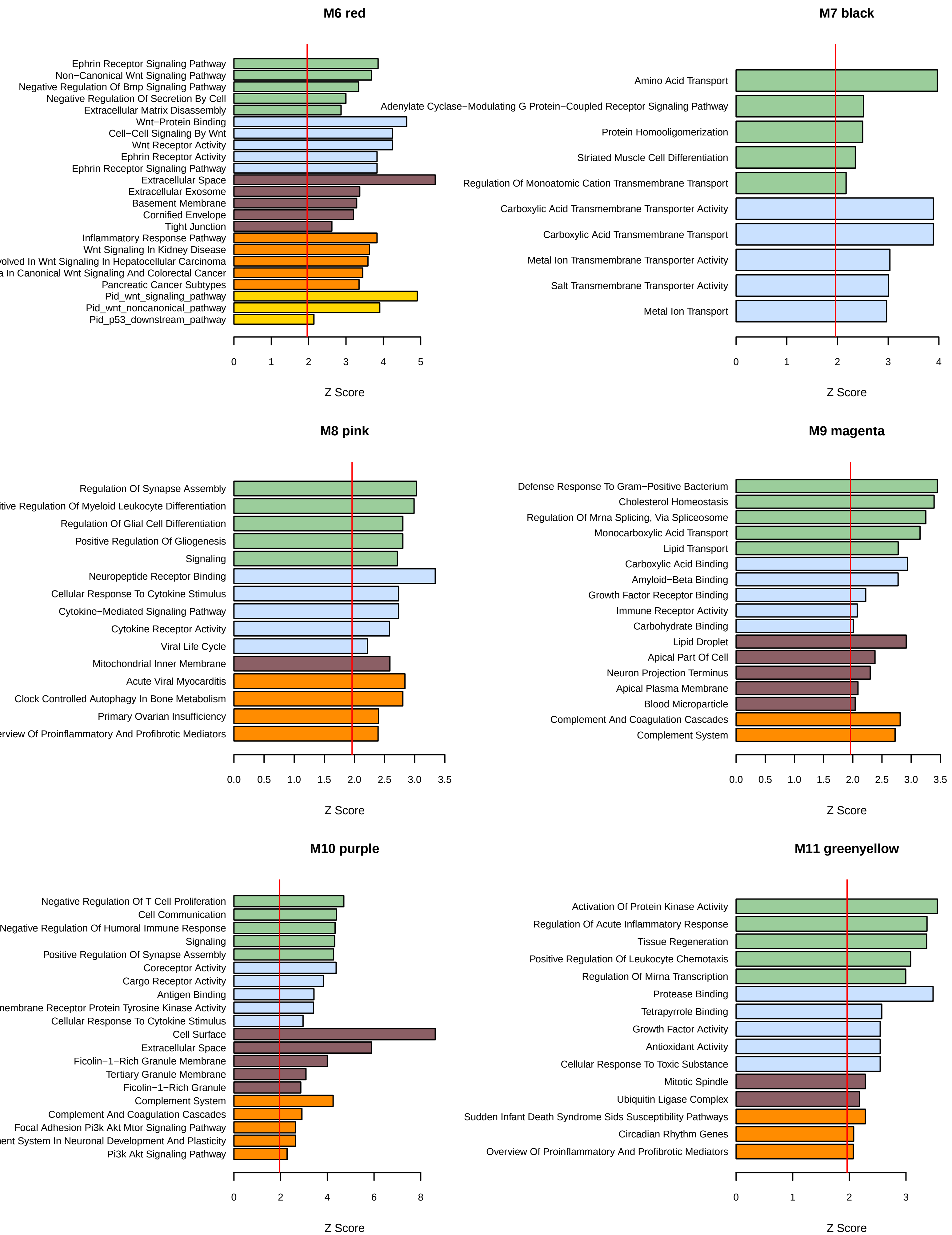

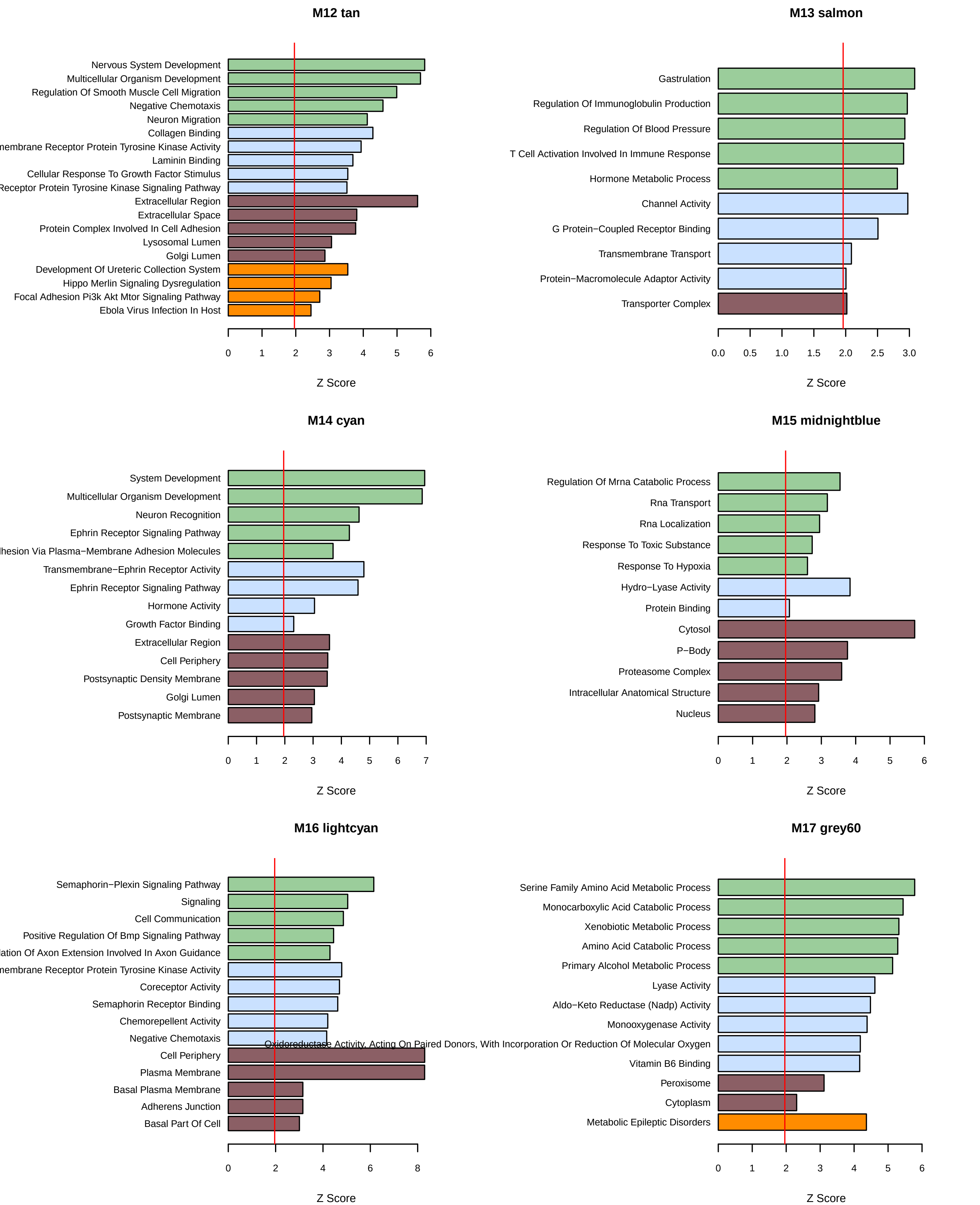

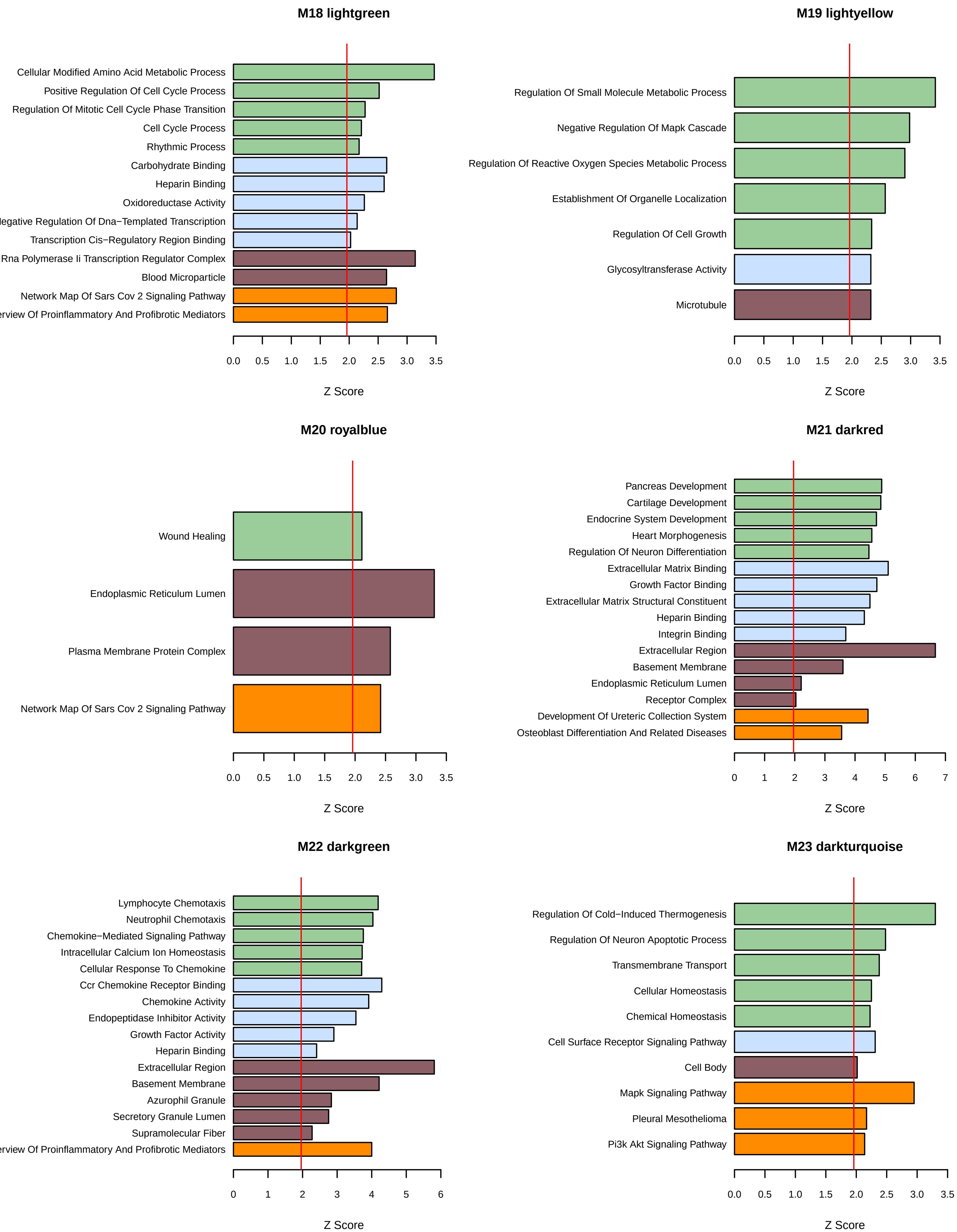

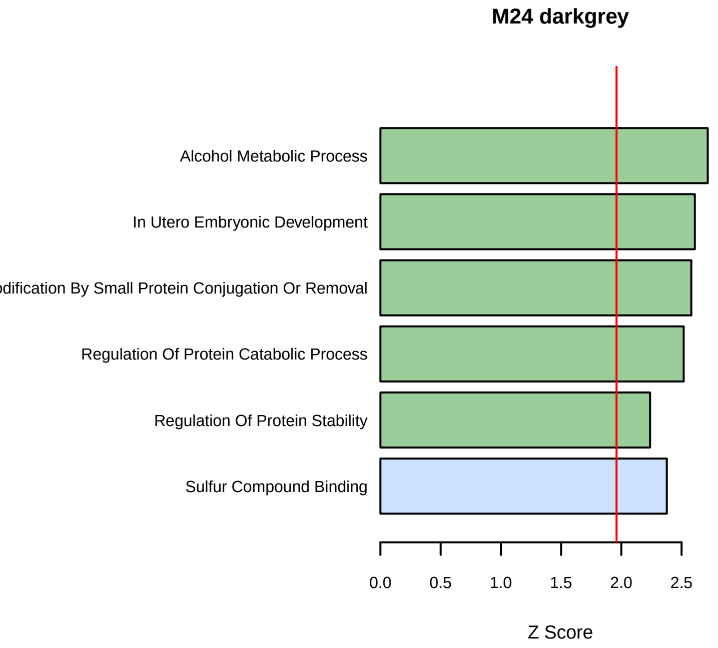

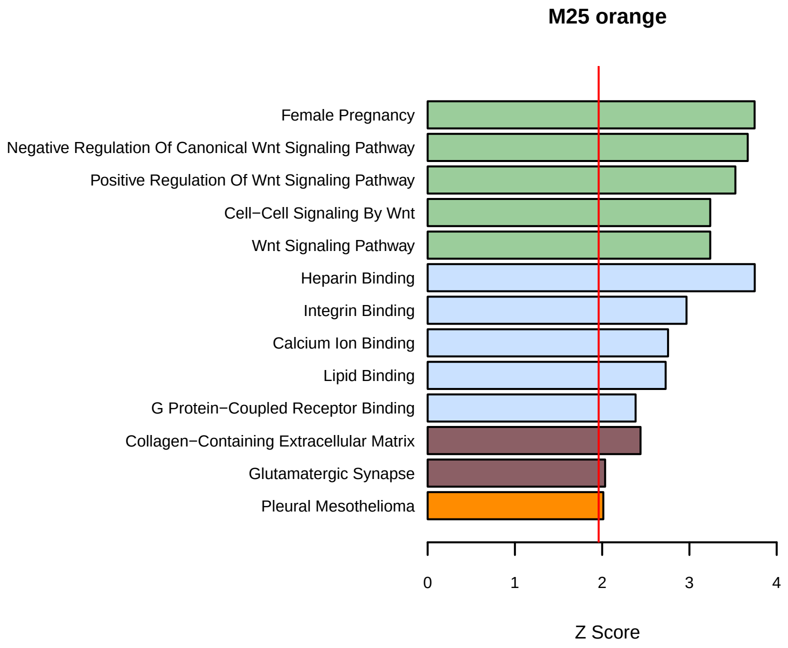

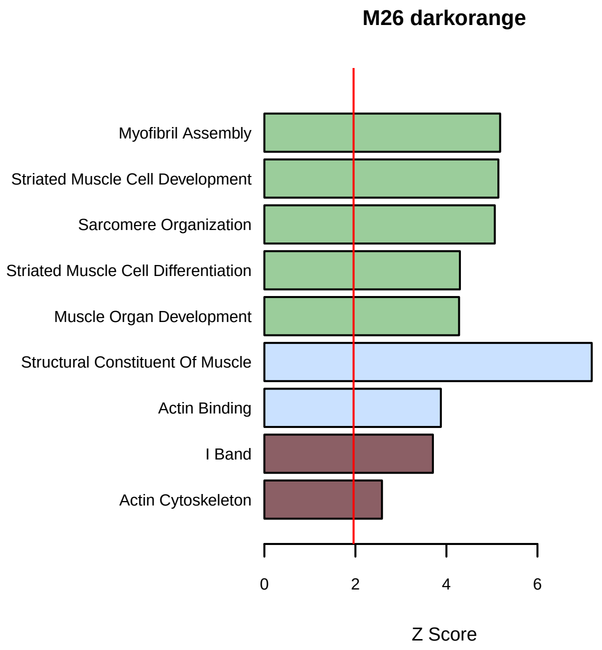

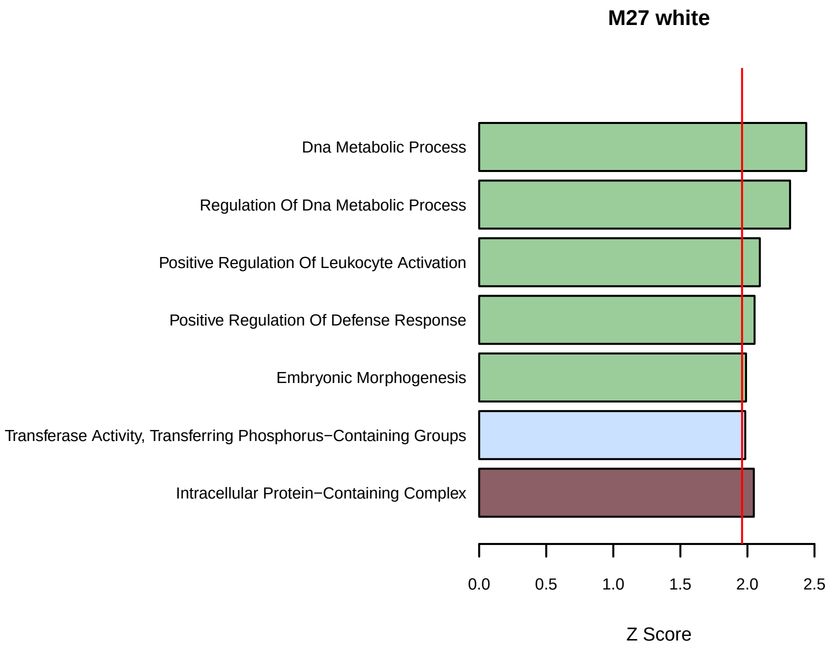

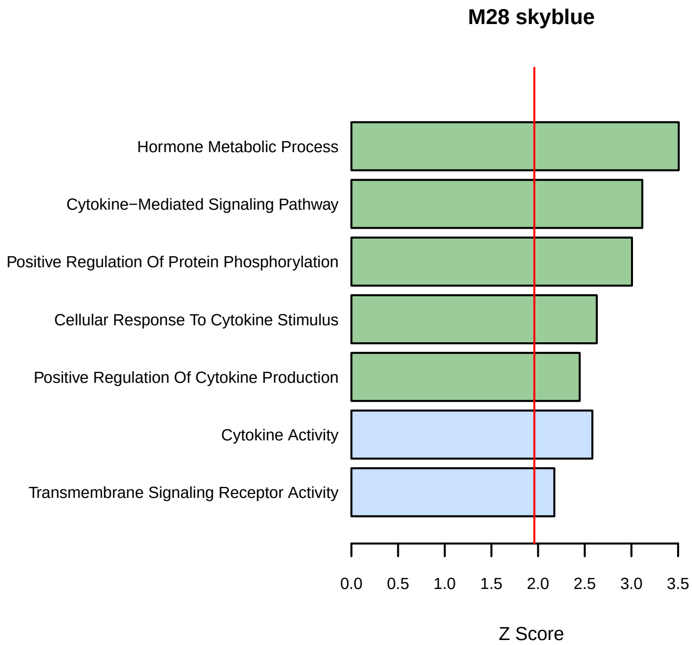

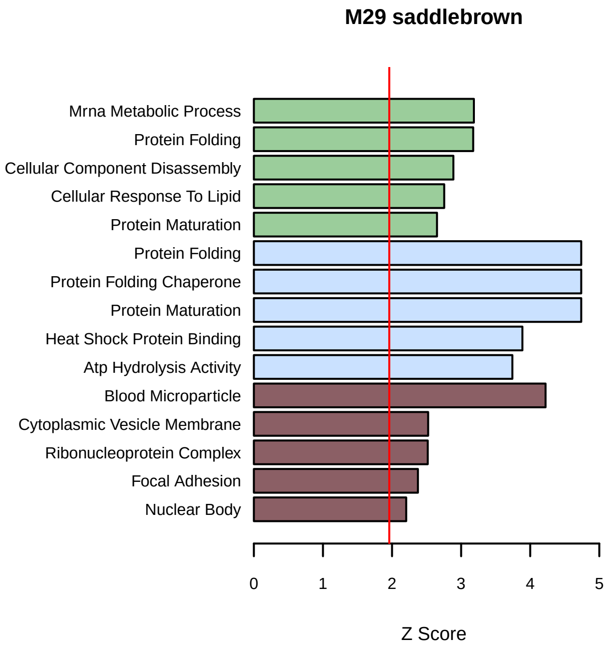

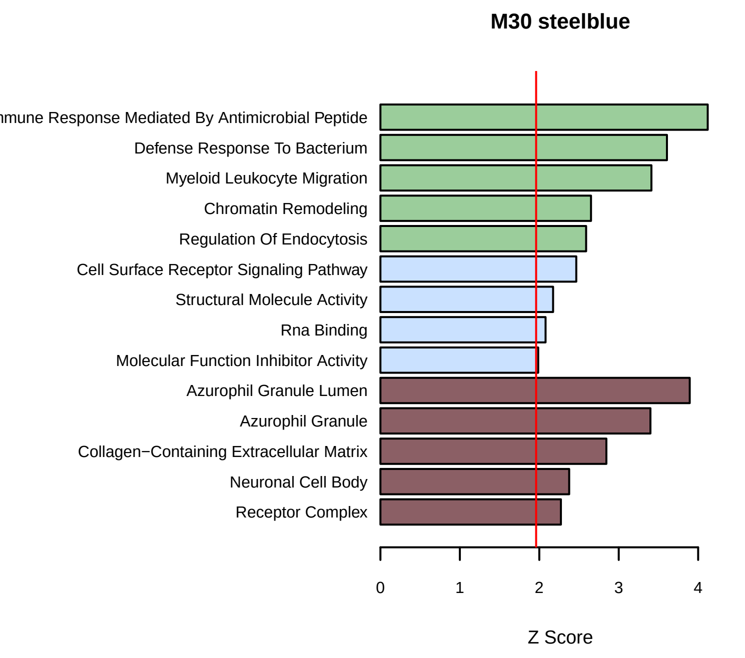

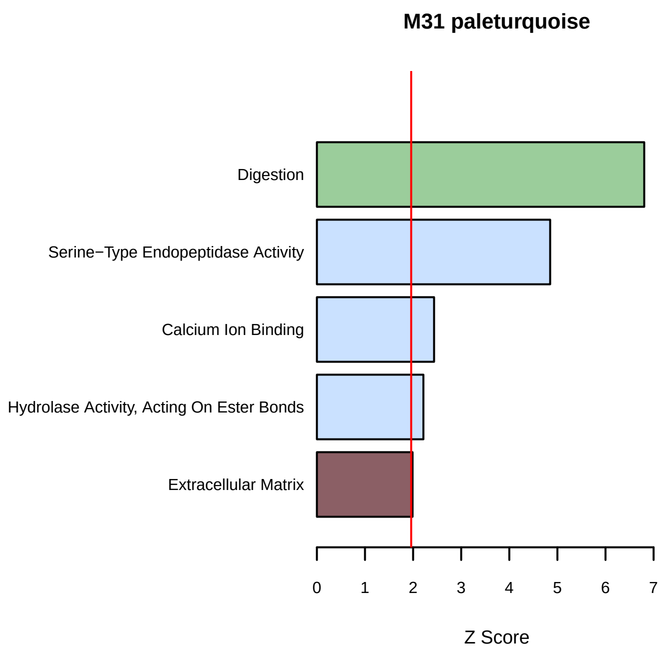

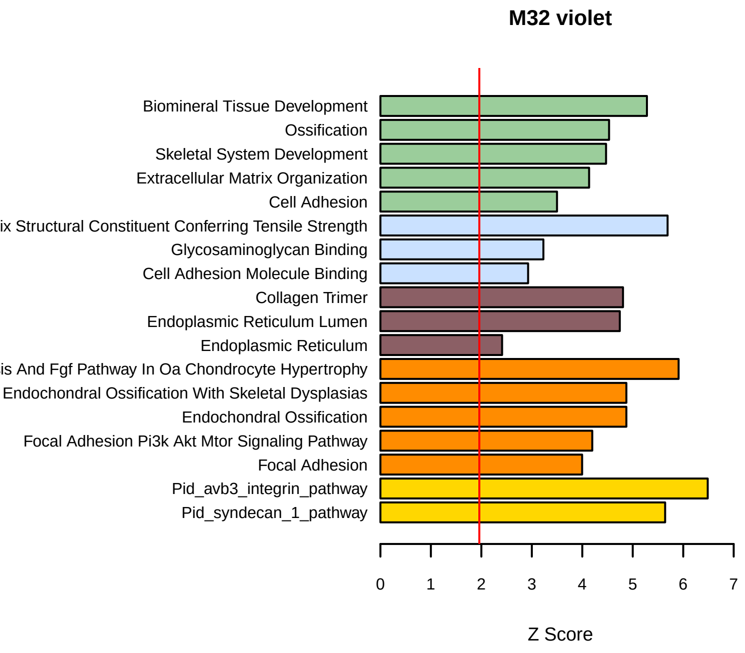
